# Deciphering Cross-Cohort Metabolic Signatures of Immune Responses and Their Implications for Disease Pathogenesis

**DOI:** 10.1101/2024.11.29.24318195

**Authors:** Jianbo Fu, Nienke van Unen, Andrei Sarlea, Nhan Nguyen, Martin Jaeger, Javier Botey-Bataller, Valerie A.C.M. Koeken, L. Charlotte de Bree, Vera P. Mourits, Simone J.C.F.M. Moorlag, Godfrey Temba, Vesla I. Kullaya, Quirijn de Mast, Leo A.B. Joosten, Cheng-Jian Xu, Mihai G. Netea, Yang Li

## Abstract

The intricate interplay between circulating metabolites and immune responses, though crucial to disease pathophysiology, remains poorly understood and underexplored in systematic research. Here, we performed a comprehensive analysis of the immune response and circulating metabolome in two Western European cohorts (534 and 324 healthy individuals) and one from sub-Saharan Africa (323 healthy donors). At metabolic level, our analysis uncovered sex differences in the correlation between phosphatidylcholine and cytokine responses upon *ex-vivo* stimulations. Notably, sphingomyelin showed a significant negative correlation with the monocyte-derived cytokine production in response to *Staphylococcus aureus* stimulation, a finding validated through functional experiments. Subsequently, employing Mendelian randomization analysis, we established a link between sphingomyelin and COVID-19 severity, providing compelling evidence for a modulatory effect of sphingomyelin on immune responses during human infection. Collectively, our results represent a unique resource (https://lab-li.ciim-hannover.de/apps/imetabomap/) for exploring metabolic signatures associated with immune function in different populations, highlighting sphingomyelin metabolism as a potential target in treating inflammatory and infectious diseases.

## Introduction

Human metabolism and immune response are closely linked, playing an important role in maintaining human health (*1*). While the immune system protect body against pathogens and maintains tissue homeostasis, these functions come at a significant bioenergetic cost, requiring precise control of cellular metabolic pathways (*2*). In turn, various metabolites serve not only as energy sources or building blocks of cellular function, but also as modulators of immune responses (*3*). An abnormal interaction between cellular metabolism and immune responses contributes to autoimmune, metabolic, and infectious diseases (*4, 5*). Pro-inflammatory cytokines are involved in promoting inflammation and modulating adaptive immune responses, which are fundamental components of the immune response. In addition, many studies have shown that pro-inflammatory cytokines, such as tumor necrosis factor (TNF) (*6*), interleukin 6 (IL-6) (*7, 8*), IL-1β (*9*), and interferon-gamma (IFN-γ) (*10*), can influence insulin resistance, adipose tissue inflammation and regulate obesity-related metabolism. On the other hand, metabolite reprogramming can modulate inflammatory states. As just a few examples, uric acid promotes IL-1β production in peripheral blood mononuclear cells (*11*), palmitate induces the secretion of IL-1β and IL-18 by macrophages (*12*), while TCA cycle metabolites such as fumarate, mevalonate and itaconate modulate trained immunity responses (*13*).

Since immunity and metabolism play crucial roles and interact in health and disease, research in the field of immunometabolism has been steadily increasing in recent years (*14–16*). However, a systematic assessment of the interplay between circulating metabolites and cytokine responses is missing, due to the difficulty to measure both immune response and metabolomic data from the same biological sources, such as identical samples and cell systems, across multiple cohorts. Furthermore, metabolic and immune interactions can vary considerably across diverse ethnic and geographical backgrounds due to genetic, environmental, dietary and lifestyle factors (*17–20*).

In this study, we analyzed plasma metabolite concentrations and cytokine responses in three distinct cohorts, totaling 1,181 individuals, including two from Western Europe and one from Sub-Saharan Africa. In total, we investigated the relationships between 4,361 metabolite features and immune cytokine responses, including 172 different cytokine production responses to stimuli. Our study aimed to understand the interaction between cytokine responses and metabolic interactions across different populations, in both men and women. To achieve this, we first analyzed the correlations between metabolite features and immune cytokine responses in each cohort. Subsequently, we conducted enrichment analyses to identify metabolites significantly associated with specific cytokine responses. We experimentally validated the relationships between specific metabolites and cytokine responses in *in-vitro* models of cytokine production stimulation assays. Subsequently, we employed Mendelian randomization (MR) analysis to explore the causal relationships between the metabolites and infectious diseases such as COVID-19, to demonstrate the importance of these interactions in actual human infections. Finally, we integrated all metabolite-cytokine association results into a publicly available database, offering insights into immunity and metabolism interactions, and supporting new treatment development.

## Results

### Circulating metabolome and innate/adaptive immune response profiling across cohorts

To comprehensively understand the relationship between metabolites and immune cytokine responses, we integrated data from two different European healthy populations: Cohort_EU1 and Cohort_EU2 from Western Europe (Netherlands) with 534 and 324 participants respectively, and one African cohort: Cohort_AF from sub-Saharan Africa (Tanzania) with 323 participants.We quantified plasma metabolite features across these cohorts using flow-injection TOF-M, identifying 1377 metabolites features in Cohort_EU1, 1373 features in Cohort_EU2, and 1611 features in Cohort_AF. In parallel, we assessed innate and adaptive immune response by measuring cytokine production in response to various stimuli: Cohort_EU1 was assessed for 6 cytokines across 18 stimulations, Cohort_EU2 for 4 cytokines across 2 stimulations, and Cohort_AF for 5 cytokines across 10 stimulations (**Fig. 1A**). **Fig. 1B** provides an overview of the study design, selecting Cohort_EU1 and Cohort_AF as discovery cohorts and Cohort_EU2 as the replication cohort.

**Fig. 1.**
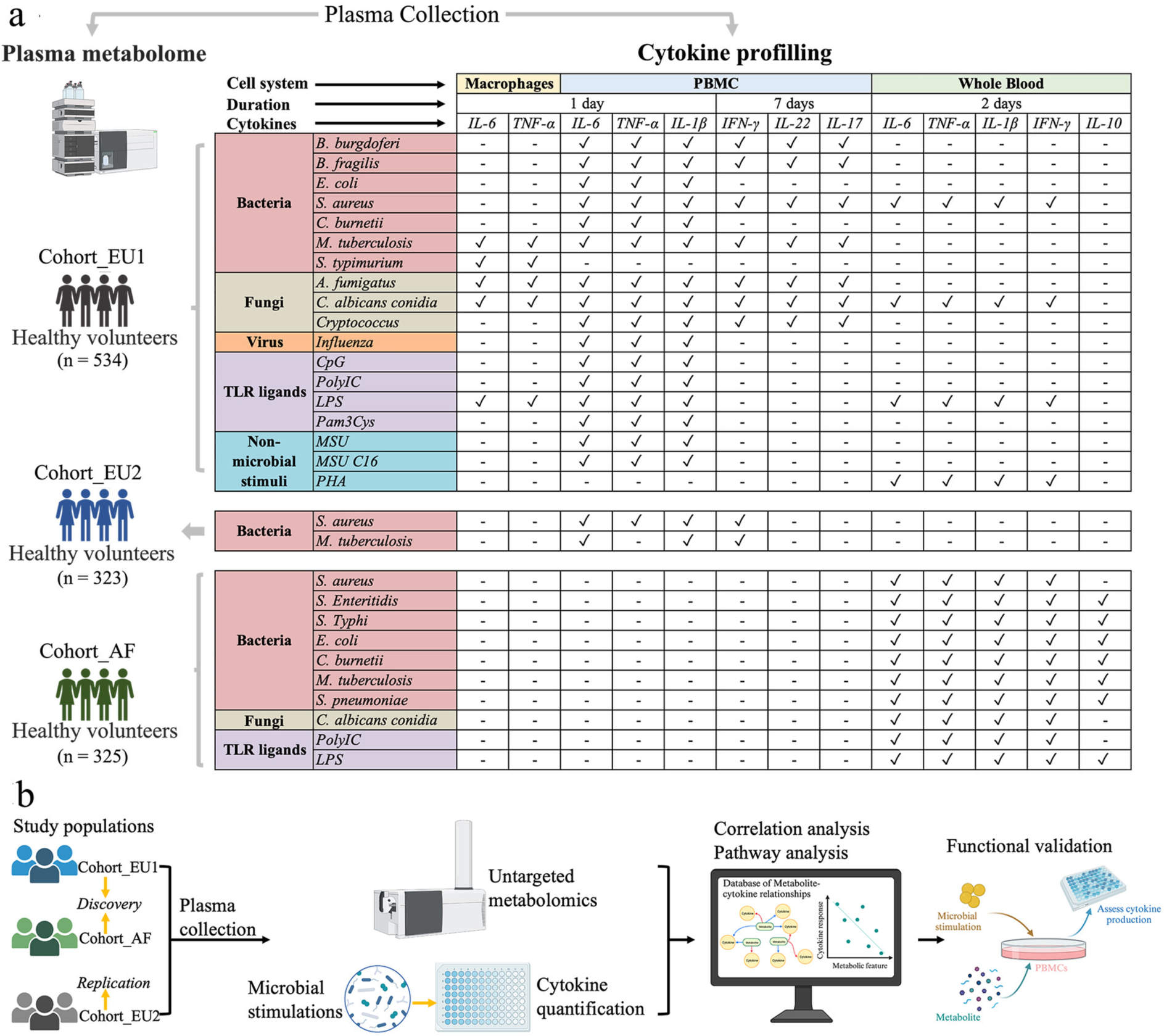
Study Overview. (a) The study includes three cohorts from the Human functional genomics project: two originating from Western Europe (Cohort_EU1 and Cohort_EU2) and one from Sub-Saharan Africa (Cohort_AF). Blood samples were collected from these cohorts for untargeted metabolomic measurement and stimulation experiments, measuring cytokine production after various human pathogens stimulation. (b) The study design initiates with the selection of cohorts for discovery (Cohort_EU1 and Cohort_AF) and replication (Cohort_EU2). To investigate the interplay between metabolic signatures and cytokine response, we conducted the correlation analysis to identify the relationships between metabolites and cytokine response. Subsequently, we performed ex vivo functional validation using PBMCs to evaluate the influence of metabolic changes on cytokine production following microbial stimulation.

### Robust plasma metabolic pathways for immune functions across European and African populations

We initiated our analysis by examining metabolic networks through Weighted Gene Co-expression Network Analysis (WGCNA) to reveal interactions between metabolism and immune responses. Specifically, we assessed metabolite co-expression networks associated with immune phenotypes (IL-1β, IL-6, TNF, and IFN-γ) following *S. aureus* stimulation, as these measurements were shared across the three cohorts. Of note, the peripheral blood mononuclear cells (PBMCs) in Cohort_EU2, the whole blood (WB) in Cohort_AF, and both WB and PBMCs in Cohort_EU1 were examined. We identified 11, 11, 10, and 10 modules of highly correlated metabolites in Cohort_EU1 (PBMCs), Cohort_EU1 (WB), Cohort_AF, and Cohort_EU2, respectively (as shown in **fig. S1A-D**, and **tables S1-3**). Subsequently, we correlated the metabolites modules with the both monocyte-derived cytokines (IL-1β, IL-6, and TNF) and T cell-derived cytokine (IFN-γ) profiles following *S. aureus* stimulation. We identified 7, 7, 8, and 7 metabolite modules that were found to be nominally significant in association with cytokine responses induced by *S. aureus* in Cohort_EU1 (PBMCs), Cohort_EU1 (WB), Cohort_AF, and Cohort_EU2, respectively, as shown in **Fig. 2A-D** (p < 0.05).

**Fig. 2.**
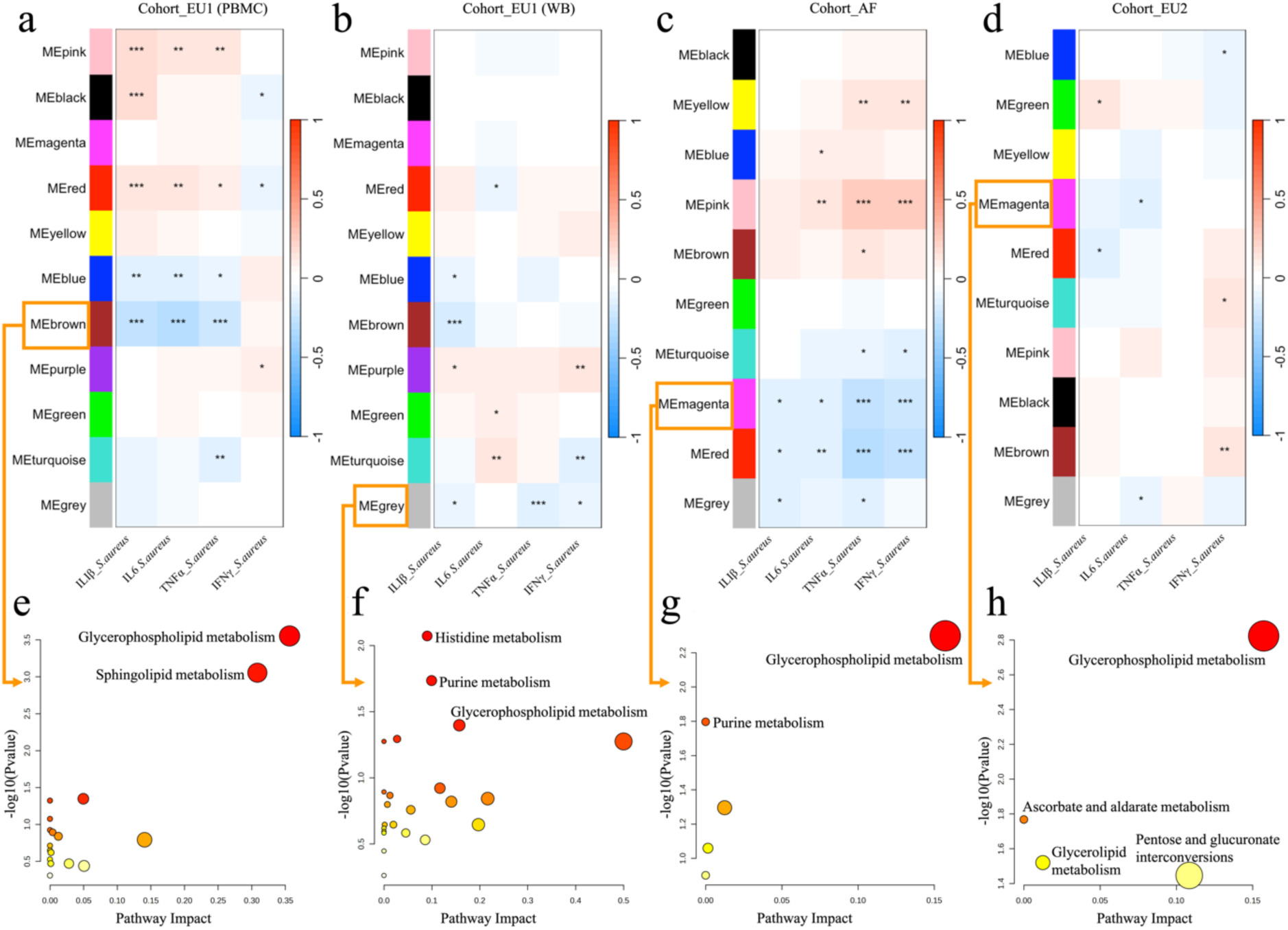
Metabolite modules associated with cytokine responses induced by S. aureus in African and European population. The specific metabolites within each module delineated by WGCNA are detailed in the Supplementary Table 1 (a, b, c and d). In three cohorts (Cohort_EU1, Cohort_AF and Cohort_EU2), 11, 11, 10 and 10 modules respectively were found to correlate with cytokine responses (IL-1β, IL-6, TNF, and IFN-γ) induced by *S. aureus* (*p<0.05, **p<0.01, ***p<0. 001). (e, f, g and h) Pathway analysis results for metabolites within the MEbrown (a), MEgrey (b), MEmagenta (c), and MEmagenta (d) modules. The darker color and larger size of the bubble dots indicate a larger -log(Pvalue), suggesting higher significance.

In total, four metabolite modules were associated with monocyte-derived cytokines (IL-1β, IL-6, TNF) in the PBMCs of Cohort_EU1 stimulated by *S. aureus*. The brown and blue modules exhibited negative correlations with these cytokine productions, while the pink and red modules showed positive correlations (**Fig. 2A**). The metabolites of the brown module were enriched in the glycerophospholipid metabolism (*p* = 0.00028) and sphingolipid metabolism pathways (*p* = 0.00088) (**Fig. 2E** and **table S4**); whereas the metabolites of the blue module were enriched in the primary bile acid biosynthesis and steroid biosynthesis pathways (**fig. S2C** and **table S4**). On the other hand, the metabolites of the pink and red modules were enriched in the pathways of nicotinate and nicotinamide metabolism, alanine, aspartate and glutamate metabolism and aminoacyl-tRNA biosynthesis, as well as vitamin B6 metabolism, tyrosine metabolism pathways (**fig. S2A-B** and **table S4**).

We also identified a grey module correlated with cytokine production (IL-1β, TNF, and IFN-γ) in the WB of Cohort_EU1 stimulated to *S. aureus* (**Fig. 2B**). Metabolites of this module were enriched in histidine metabolism (*p* = 0.00846), purine metabolism (*p* = 0.01840) and glycerophospholipid metabolism (*p* = 0.04006, **Fig. 2F** and **table S4**). In Cohort_AF, we observed two metabolite modules (magenta and red) negatively correlated with both monocyte-derived cytokines (IL-1β, IL-6, TNF) and T cell-derived cytokine (IFN-γ) production after *S. aureus* stimulation (**Fig. 2C**). Metabolites of the magenta module was enriched in glycerophospholipid metabolism (*p* = 0.00502) and purine metabolism (*p* = 0.01596, **Fig. 2G** and **table S5**), while metabolites of the red module was enriched in pyrimidine metabolism, nicotinate and nicotinamide metabolism, and lysine degradation (**fig. S2D** and **table S5**). It is worth noting that the glycerophospholipid metabolism pathway was consistently identified in both Cohort_EU1 (PBMCs and WB) and Cohort_AF, underscoring its robustness (as shown in **Fig. 2E-G**). This result was further validated in the replication cohort (Corhort_EU2). The magenta module, negatively correlated with IL-6, showed enrichment in the glycerophospholipid metabolism pathway (**Fig. 2D, 2H**, **S3** and **table S6**). This finding aligns with the known role of glycerophospholipid metabolism is known to play an important role in regulating immune responses (*21, 22*).

**Fig. 3.**
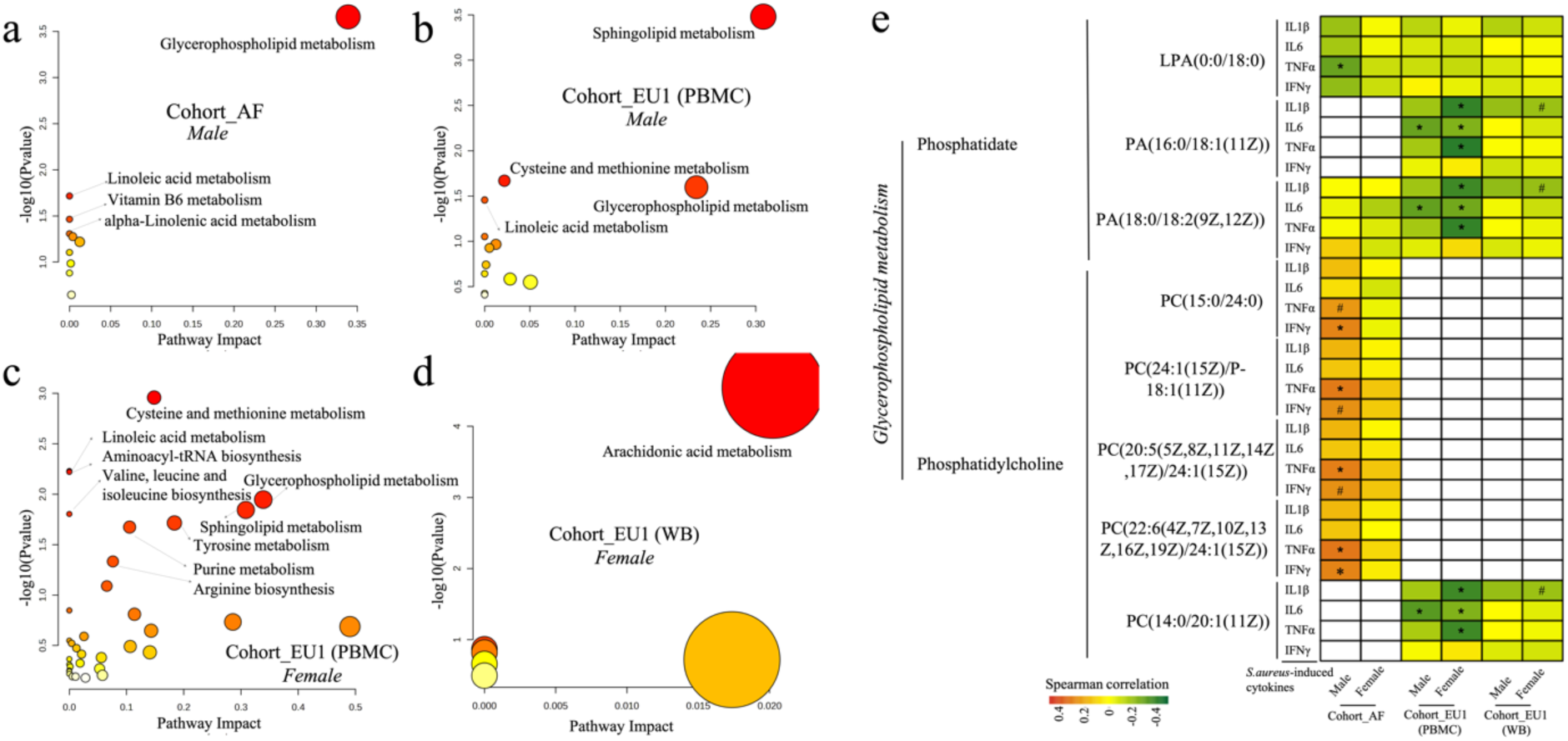
Impact of sex differences on the correlation between metabolites and cytokine responses. Scatter plots illustrate pathway analysis results for significant correlations (FDR < 0.05) between metabolites and cytokine responses, with distinctions based on gender in varied cohorts: (a) Cohort_AF, male; (b) Cohort_EU1 (PBMCs), male; (c) Cohort_EU1 (PBMCs), female; (d) Cohort_EU1 (WB), female. The darker the bubble color, the larger the -log(Pvalue), indicating higher significance. (e, f) Differences in the Glycerphospholipid and Linoleic acid metabolism between males and females. Red signifies a significant positive correlation between the metabolite and cytokine response, while green indicates a significant negative correlation. Cells shaded in light red and light green represent 0.05 < FDR < 0.10, with light red suggesting a trend towards positive correlation and light green pointing to a trend toward negative correlation. Asterisk *(FDR < 0.05) and hash # (0.05 < FDR < 0.10).

In summary, our study identified a common link between metabolic pathways and immune responses across different cohorts. Notably, glycerophospholipid metabolism emerged as a consistent pathway linked to immune regulation, underscoring its importance in the context of infection-induced immune responses.

### Sex-specific metabolic markers for immune functions

Since sex impacts both cytokine productions (*23*) and metabolic regulation (*24*), we investigated its impact on the relationship between metabolome and *S. aureus*-induced cytokine responses (IL-1β, IL-6, TNF, and IFN-γ). We had a balanced sex distribution among participants, with 50.77%, 56.33%, and 56.70% females, in Cohort_AF, Cohort_EU1, and Cohort_EU2, respectively (**fig. S2A-C**). In Cohort_EU1 (PBMCs), we found 95 metabolites in males and 302 metabolites in females that were significantly correlated with at least one cytokine response, after adjusting for the effects of age and body mass index (BMI); In Cohort_EU1 (WB), 7 metabolites in males and 52 metabolites in females showed significantly correlations with at least one cytokine response. However, no metabolites in Cohort_EU2 exhibited such correlations, possibly due to the sample size and large variability within the metabolite and cytokine data, limiting the statistical power to detect such correlations. In Cohort_AF, there were 59 metabolites in males and 10 in females with significant correlations (Spearman correlation, FDR < 0.05, **fig. S4D** and **tables S7-9**).

Interestingly, metabolites associated with cytokine response in males were prominently linked to glycerophospholipid metabolism and sphingolipid metabolism (**Fig. 3A-B**). In contrast, metabolites associated with cytokine response in females (**Fig. 3C-D**) were involved in a diverse range of metabolic pathways including cysteine and methionine metabolism, linoleic acid metabolism and aminoacyl-tRNA biosynthesis, with arachidonic acid metabolism having particularly notable effects. Pathway analysis is constrained by the limited number of significant associations. Despite these limitations, sex differences in the major metabolic pathways were observed, with some shared pathways such as sphingolipid metabolism associated with cytokine response in both male and female subjects.

In addition, the metabolites from the glycerophospholipid metabolism pathway were associated with cytokine response in both males and females across different cohorts (**Fig. 3A-C** and **fig. S5A-D**). Within the glycerophospholipid metabolism pathway, phosphatidate metabolites were generally negatively correlated with monocyte-derived cytokines responses, while phosphatidylcholine metabolites displayed variable correlations with cytokines responses across different populations (**Fig. 3E**).

Among phosphatidate metabolites, lysophosphatidic acid (LPA)(0:0/18:0) showed a significant negative correlation with TNF response in males of the Cohort_AF, whereas both phosphatidic acid (PA)(16:0/18:1(11Z)) and PA(18:0/18:2(9Z,12Z)) displayed significant negative correlations with *IL-6* response in both males and females of Cohort_EU1 (PBMCs). Furthermore, PA(16:0/18:1(11Z)) and PA(18:0/18:2(9Z,12Z)) also exhibited significant negative correlations with IL-1β and TNF response in females of the Cohort_EU1(PBMCs) and Cohort_EU1(WB).

In the phosphatidylcholine group of the glycerophospholipid metabolism pathway, a positive correlation trend with TNF and IFN-γ response was observed solely in males of the Cohort_AF. By contrast, this phosphatidylcholine group showed negative correlations with cytokine response for both males and females in the European cohorts. For instance, phosphatidylcholine (PC)(14:0/20:1(11Z)) displayed a significant negative correlation with IL-6 in both males and females in Cohort_EU1 (PBMCs). Additionally, this metabolite showed negative correlations with IL-1β and TNF response in females in cohort_EU1 (PBMCs) and a negative correlation trend with IL-1β in females in Cohort_EU1 (WB).

In summary, metabolites such as phosphatidylcholine can serve as sex-specific metabolic markers for innate immune responses, aligning with previous studies highlighting sex disparities in these metabolites (*25*). This underscores the importance of considering sex disparities in the contribution of the glycerophospholipid metabolites to immune function, essential for both research and therapeutic interventions.

In addition to identifying the glycerophospholipid metabolism pathway, we also discovered the regulatory role of the sphingomyelin metabolism pathway on immune responses in both males and females of cohort_EU1 (PBMC) (**Fig. 3B-C**), a pathway known for its key role in immune regulation (*26*). Moreover, in females, metabolites associated with cytokine response were significantly enriched in arachidonic acid metabolism (**Fig. 3D**). Previous studies have highlighted the relevance of arachidonic acid metabolism to the presence of sex differences (*27, 28*).

Altogether, these findings underscore the intricate interplay between metabolite and cytokine response across sex and various ethnicities.

### Sphingolipid metabolism consistently correlates with the capacity of monocyte-derived cytokine production

Next, we investigated the relationship between individual metabolic features and immune responses to *S. aureus*. In total, we identified 255, 423, and 167 metabolites in Cohort_AF, Cohort_EU1 (PBMCs), and Cohort_EU1 (WB), respectively, significantly correlated with at least one cytokine response, as shown in **fig. S6A** and **tables S10-12** (FDR < 0.05). No metabolites in Cohort_EU2 showed such correlations, possibly due to insufficient sample size reducing statistical power, leading to an inability to detect smaller but potentially meaningful biological effects after FDR correction for multiple comparisons. Based on pathway analysis, the citrate cycle (TCA cycle) and sphingolipid metabolism stood out as the most statistically significant pathways within the Cohort_AF (**Fig. 4A**). While there was a more diverse set of pathways in Cohort_EU1 using PBMC samples (**Fig. 4C**) compared to WB samples (**Fig. 4B**), sphingolipid metabolism was also among the top significant pathways in both two cohorts. Thus, sphingolipid metabolism emerges as a common signature across all cohorts, suggesting its universal importance in the context of metabolite-cytokine correlations. This finding aligns with the known role of sphingolipid metabolism and its derived metabolites in immune responses (*26, 29, 30*). Specifically, in Cohort_AF, monocyte-derived cytokines, such as IL-6 and TNF exhibited a negative correlation with SM, whereas IFN-γ and IL-1β showed minimal correlation (**fig. S7**). In Cohort_EU1 (WB), a significant negative correlation between the monocyte cytokine IL-1β and SM (p = 2.6e-05) was found, but not for IFN-γ response (**fig. S7-8**). Similarly, in Cohort_EU1 (PBMCs) SM displayed a consistent negative correlation with monocyte-derived cytokines like IL-6, IL-1β, TNF, but not with IFN-γ response (**fig. S7-8**). In Cohort_EU2, IL-6 was negatively correlated with SM (**fig. S7**).

**Fig. 4.**
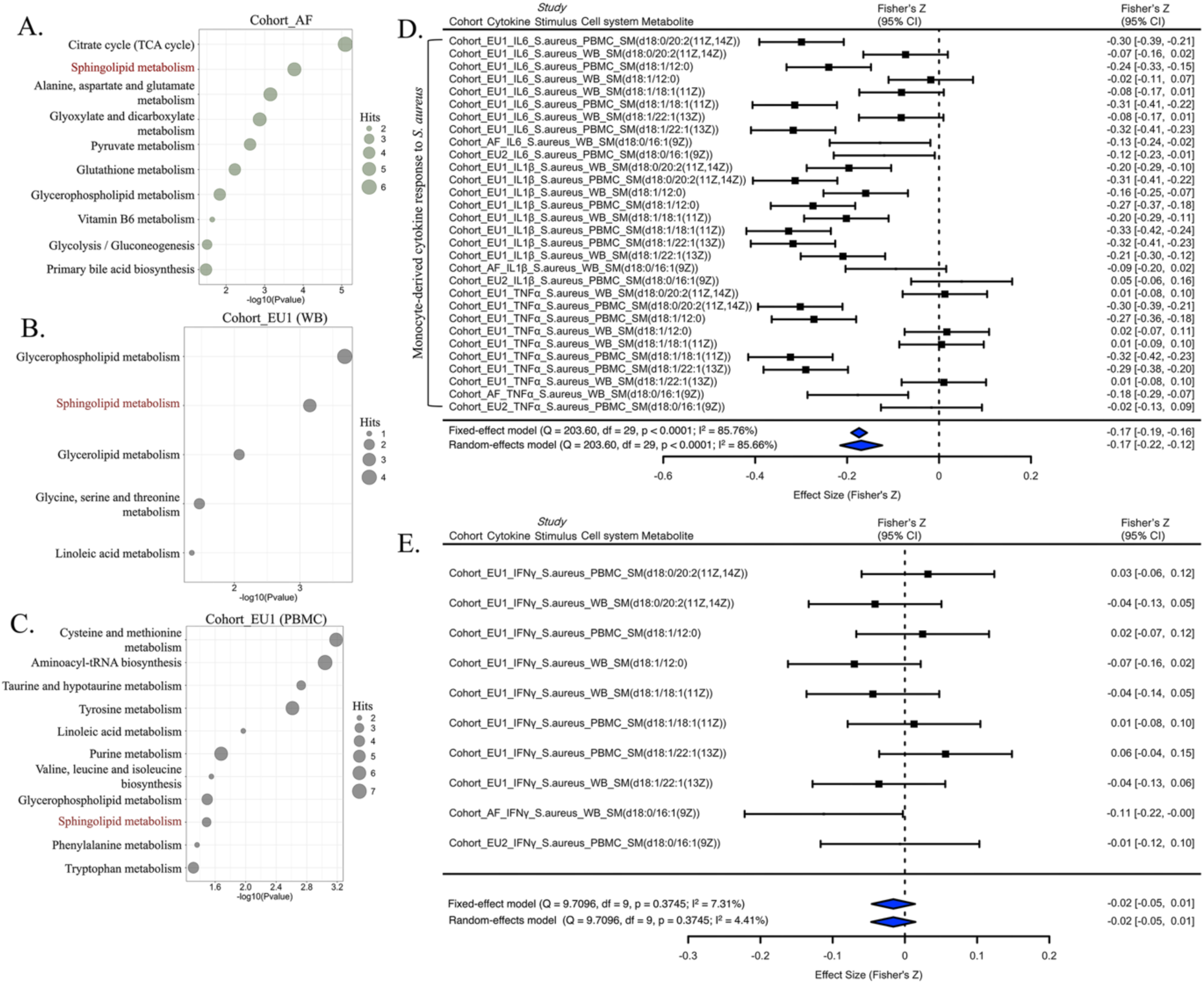
Immune response-related metabolites across multiple cohorts. Bubble plots showing the results of pathway analysis of metabolites significantly correlated (FDR < 0.05) with S. aureus-induced cytokine responses (IL-1β, IL-6, TNF, and IFN-γ) in Cohort_AF (a), Cohort_EU1 (b), and Cohort_EU1 (PBMCs) (c). (d and e) A ’forest’ plot displaying different mean values (center of symbols), confidence limits (95% confidence intervals), and precision levels (denoted by the size or ’weight’ of the symbols, where larger symbols signify higher precision). These are shown for the effect sizes derived from individual studies (in black), as well as the aggregate mean values (center of symbols) and 95% confidence intervals (width of symbols) calculated through meta-analysis using both a fixed-effect model and a random-effects model (both in blue).

Given the above indications that SM may be more closely related to monocyte-derived cytokines rather than IFN-γ, we then performed the meta-analysis of the correlation coefficients between SM and *S. aureus*-induced cytokine production in three cohorts. Here, we have used both the fixed-effect model and random-effect model for computation. The results (**Fig. 4D**) all showed that circulating SM is significantly negatively correlated with monocyte-derived cytokines (IL-6, TNF, and IL-1β) (Fixed-effect model: pooled r = -0.1740, 95% CI -0.1913 — -0.1567, *p* < 0.0001; random-effect model: pooled r = -0.1694, 95% CI -0.2151 — -0.1236, *p* < 0.0001). Additionally, there is no correlation between circulating SM and T cell-derived cytokine (IFN-γ) response (Fixed-effect model: pooled r = -0.0159, 95% CI -0.0459 — 0.0140, *p* = 0.2969; random-effect model: pooled r = -0.0160, 95% CI -0.0467 — 0.0146, *p* = 0.3052) (**Fig. 4E**).

In summary, SM showed consistent negative correlations with monocyte-derived cytokine responses in the multiple cohorts, suggesting a potential inhibitory effect of SM on the innate immune function. To validate this funding, we stimulated peripheral blood mononuclear cells (PBMCs) with either S. aureus or lipopolysaccharide (LPS) for 24h, followed by measurement of proinflammatory cytokines in the supernatant of the stimulated cells (**Fig. 5**). Two different sources of sphingomyelin (chicken yolk and porcine brain) were used, and both showed 30% to 50% inhibition of TNF and IL-1β production in a dose-dependent manner (**Fig. 5A-B**). In contrast, the induction of IL-6 by the two microbial stimuli was less strongly inhibited by sphingomyelin (**Fig. 5C**).

**Fig. 5.**
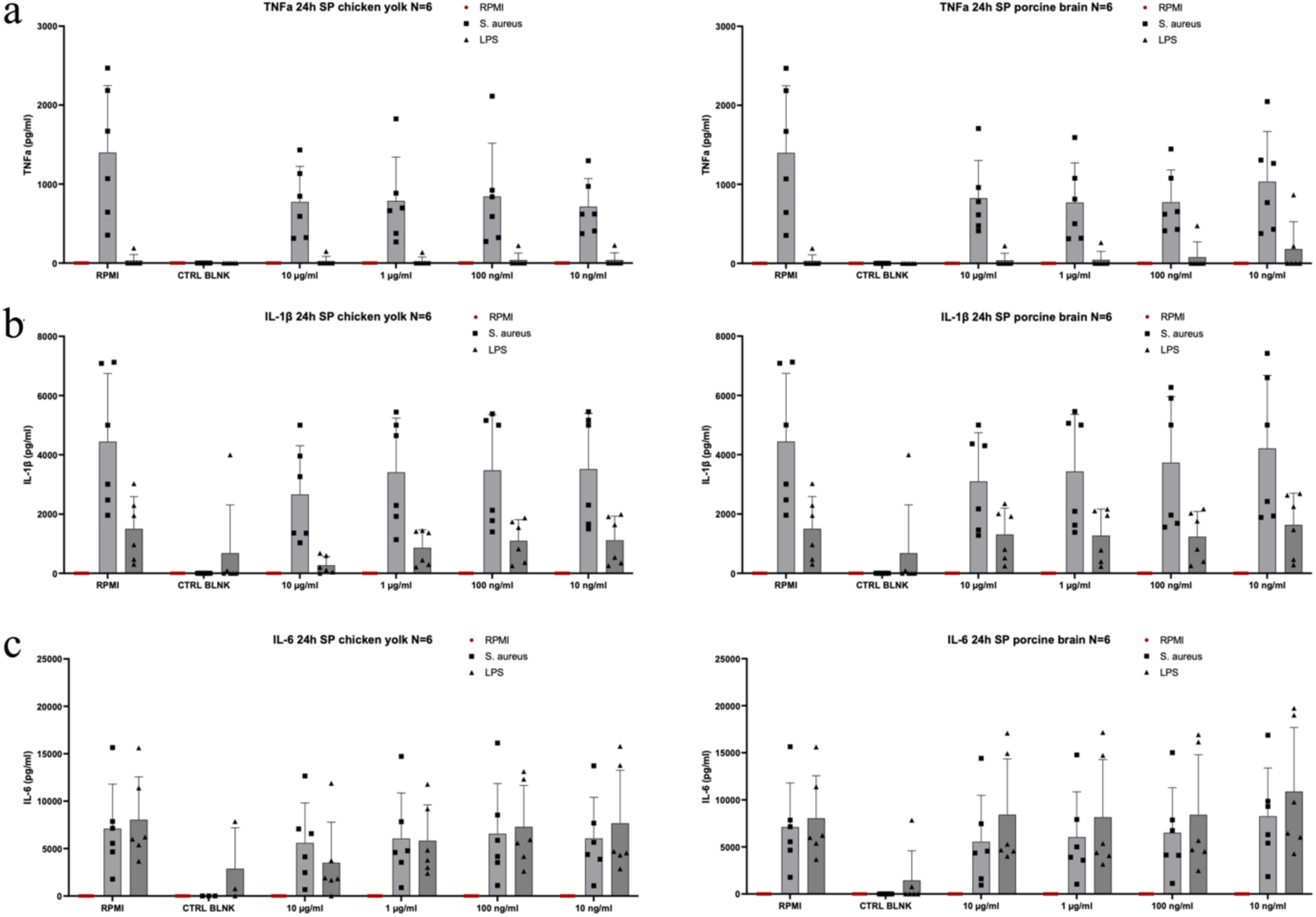
Human PBMCs were stimulated with LPS or heat-killed *S*. aureus in the absence of presence of chicken yolk or porcine brain sphingomyelin. TNF (a), IL-1β (b) and IL-6 (c) were measured in the supernatants of the stimulated PBMCs by ELISA. Experiments were performed in two independent experiments with a total of six volunteers.

### Genetic analysis shows the causal role of Sphingomyelins in COVID-19

The results described above suggests an inhibitory role of SM metabolite on innate immune function. We next examined whether SM could be a potential modulator for cytokine responses and may hold potential therapeutic significance in diseases, such as managing the cytokine overproduction in COVID-19 patients that can lead to intense inflammation, organ damage, and even death (*31*). Firstly, we used a public metabolomics dataset comprising 198 individuals with COVID-19 of varying severities (*32*) to examine the association between all sphingomyelin metabolites and the disease severity. The concentrations of sphingomyelins varied among healthy donors and different severity groups of COVID-19 patients: mild, moderate, and severe (**fig. S9**). In general, the concentration of the majority of SM metabolites (**fig. S9**) was significantly higher in the healthy donors compared to the COVID-19 patients, and negatively associated with COVID-19 severity. For several SM metabolites, such as SM (d18:0/18:0, d19:0/17:0), SM (d18:1/20:2, d18:2/20:1, d16:1/22:2), SM (d18:1/22:2, d18:1/22:1, d16:1/24:2), and SM (d18:2/24:2) (**fig. S9 I**, **L**, **Q** and **U**, respectively), patients with moderate symptoms showed a significant higher in SM concentrations compared to the healthy control group.

We next examined the causal relationship between sphingomyelin metabolites and COVID-19 using the previously reported SM associated variants (mQTLs, or metabolic quantitative trait loci) (*33*) and the public GWAS summary statistics of COVID-19 (*34*), using Mendelian randomization (*35*) (MR) method. Using 10 independent SNPs (*p* < 5.0 × 10^−7^ and clumping variants with linkage disequilibrium *r^2^* < 0.001) as instruments, the results of two commonly used MR methods, i.e. weighted median estimator and inverse-variance weighted (*36, 37*), consistently showed that a decrease in circulating sphingomyelin concentration had a causal effect on COVID-19 severity (*p* = 4.79 × 10^−2^, and 7.92 × 10^−3^, respectively; effect sizes = −0.12, and −0.13, respectively; **Fig. 6A-B**, **table S13**). A forest plot of the 10 plasma SM SNPs associated with the risk of COVID-19 is shown in **Fig. 6C**. The plot shows each SNP’s effect size and 95% Confidence Interval (CI) with points and lines; the negative estimates suggest that higher SM concentrations decrease the risk of developing severe COVID-19 symptom.

**Fig. 6.**
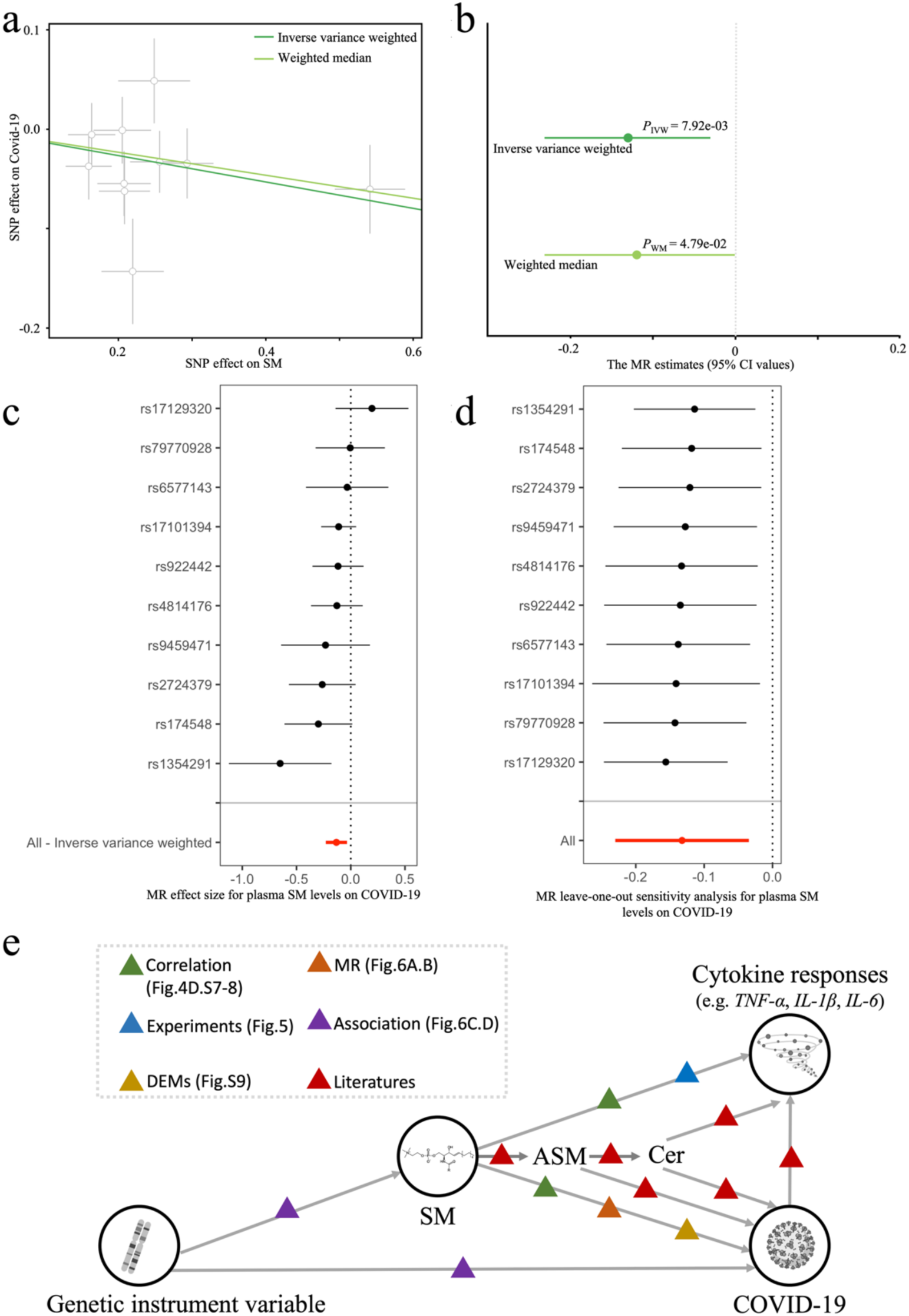
Causal relationships between sphingomyelin and COVID-19 as assessed by MR analysis. (a) The regression lines for the inverse variance weighted (IVW), weighted median (WM) method is shown. (b) Forest plot of the effects of SM on COVID-19. It shows the MR effect size (center dot) and 95% CI, estimated with the IVW MR approach. (c) Forest plot of the 10 plasma SM SNPs associated with risk of COVID-19. (d) Forest plots of MR leave-one-out sensitivity results. (e) A graphic summary of the regulation network of sphingomyelin, cytokines responses, and COVID-19.

Considering the heterogeneity of SNP effects, we went further to apply an aggregated approach, the Inverse Variance Weighted (IVW) method. As the red line indicates, there is a collective trend where higher SM levels correspond to a lower risk of developing severe COVID-19, providing a more reliable overall estimate. To validate the adherence of our data to MR assumptions, we conducted a series of sensitivity analyses. These included tests for horizontal pleiotropy, as indicated by a non-significant MR-Egger intercept (*p* > 0.05, detailed in **table S14**), assessments of heterogeneity via Cochran’s Q test (*p* > 0.05, shown in **table S15**), and a leave-one-out analysis (illustrated in **Fig. 6D**). We refrained from using causal estimates obtained through the MR-Egger method as a filtering criterion due to its relatively low power in detecting causality (*38*). To sum up, the robustness of the MR causal estimates was further substantiated by these sensitivity analyses.

In line with our observation of the negative correlation between SM and COVID-19 severity, previous research indicated a significant decrease in sphingomyelin (SM) concentrations and a notable increase in ceramide (Cer) concentrations in severe COVID-19 patients (*39*). Previous studies have also reported that the SARS-CoV-2 virus can activate acid sphingomyelinase (ASM) and the Sphingomyelinase-Ceramide pathway (*40*), and Cer can promote increased cytokine secretion (*41–43*). We therefore propose that the interactions between sphingomyelin (SM), enzymes of the Sphingomyelinase-Ceramide pathway, and Cer, modulate cytokine secretion and disease severity in COVID-19 patients (**Fig. 6E**). This regulatory framework connects genomic variations to COVID-19 diseases by modulating gene expression, metabolite profiles, and immune responses. This framework is constructed using multi-omics data sourced from various cohorts, public repositories, and existing literature.

### An online tool for exploring metabolic features for immune functions in human

To aid in exploring the intricate connections between metabolite features and immune function, we established the IMetaboMap online tool (https://lab-li.ciim-hannover.de/apps/imetabomap/), a pioneering resource that catalogs interactions between plasma metabolites and cytokine responses to various stimuli in humans. This tool will prove instrumental in examining how these interactions differ among various populations, between sexes, and across different tissue types. Our analysis showed that Cohort_EU1 had 125,307 metabolite-cytokine connections, while Cohort_EU2 and Cohort_AF yielded 28,833 and 80,550 connections, respectively, totaling 234,690 unique connections recorded in IMetaboMap. This tool stands as a testament to our comprehensive approach, enabling detailed exploration of the dynamic relationships between metabolites and cytokines, and shedding light on specific associations that may vary by sex and population. This extensive mapping effort is visualized in **Fig. 7**, which serves as a foundational reference for ongoing and future research into the mechanisms and functions of immunometabolism, advancing our understanding of immune-metabolic interplay and paving the way for effective therapeutic interventions.

**Fig. 7.**
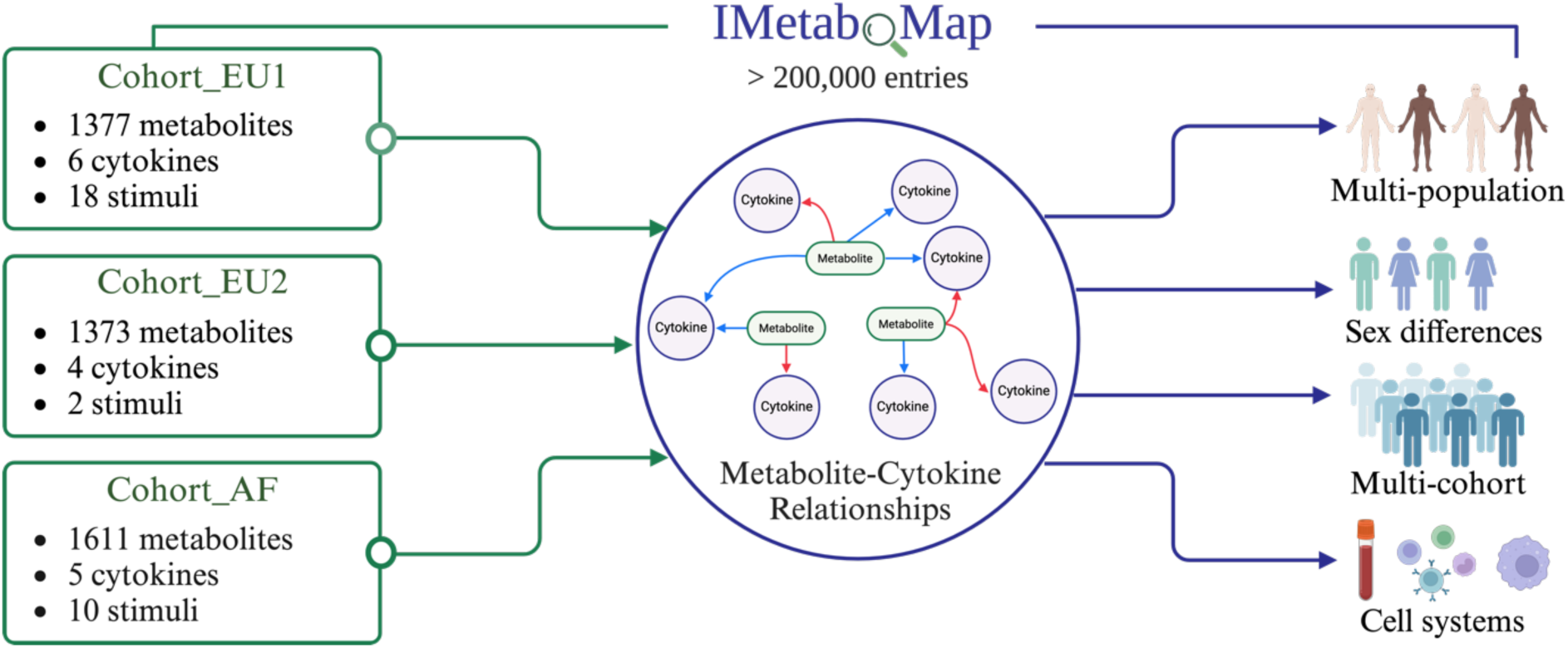
IMetaboMap is a web-based platform designed for users to explore correlations between metabolites and cytokine responses interactions. This pioneering tool offers insights into the interplay between plasma metabolites and cytokine responses to various stimuli in humans, with the capability to analyze variations across population, sex, and cell system.

## Discussion

In this study, we conducted an in-depth analysis of the interplay between plasma metabolite features and cytokine response to various stimuli across multiple cohorts with diverse ethnicities including two Western European cohorts and one East-African cohort from Tanzania. An online tool, IMetaboMap, has been developed to offer insights into the interrelationships between metabolites and cytokine responses across different populations and sexes. Furthermore, we particularly highlighted the pivotal role of baseline metabolites in immune modulation and COVID-19.

Benefiting from the uniqueness of the data from multiple cohorts with different populations, we systematically explored the relationship between metabolites and immune phenotypes. First, we identified glycerophospholipid metabolism, a common key metabolic pathway, across different populations, to play an important role in regulating immune responses and cytokines production by human circulating immune cells (*44, 45*). Further, in the analysis of sex differences, we found that phosphatidylcholine, an important component of the glycerophospholipid metabolic pathway, can serve as a sex-specific metabolic marker, which is consistent with previous studies on sex differences in these metabolites (*25*). These findings not only provide valuable evidence for the study of the glycerophospholipid metabolic pathway, but also indicate potential targets for the study of immune related diseases.

An important finding highlights the importance of the sphingolipid metabolism pathway in the regulation of immune responses, as well as the potential of sphingolipids in modulating various immune responses and their therapeutic implications. In particular, the concentration of sphingomyelins significantly correlates with monocyte-derived cytokines response (TNF, IL-1β, IL-6), but not with T cell-derived cytokines response (IFN-γ) to *S. aureus* stimulation (**Fig. 4D-E**). This implies a stronger role for sphingomyelins in the modulation of innate immune responses rather than in the adaptive responses. The mechanism by which sphingomyelin influences immune responses are likely complex, involving cellular processes such as the cell signaling (*46*), membrane fluidity (*47*), and intercellular interactions (*48*). The role of sphingomyelin in the modulation of innate immunity, as observed in our study, might be due to its localization within cell membranes and its participation in the immediate cellular response upon recognition of pathogen-and danger-associated molecular patterns. It also suggests that sphingomyelin predominantly participates in immediate and ubiquitous defense mechanisms, such as inflammation or rapid responses to pathogens, rather than specialized, long-term defense strategies involving memory lymphocytes and antibodies. The sphingomyelin – cytokines response interaction described here supports previous studies which suggested that cytokines released by innate immune cells are more strongly influenced by circulating metabolites (*16, 49*). Thus, sphingomyelin metabolites could be potential targets for therapy in inflammation-mediated diseases.

Our study has several important strengths. First, the complementary, but independent, identification of metabolite-immune interaction in omics studies, functional experiments, and genetic Mendelian randomization provides very strong arguments for the validity of our conclusions. Second, our study proved the relevance of these findings by the identification of an association between sphingomyelin concentrations and COVID-19 severity. Whereas the significant mortality associated with COVID-19 is closely linked to the “cytokine storm” (*31*), our findings indicate an inverse relationship between elevated SM concentrations and the severity of COVID-19 infection. This suggests that modulating SM levels might regulate cytokine production, attenuate inflammatory responses, and potentially reduce mortality. In line with our findings, the presence of sphingolipids in the milk fat globule membrane has demonstrated anti-bacterial properties against several microorganisms (*50*), while milk sphingomyelin could mitigate LPS-induced inflammation in RAW264.7 macrophages (*51*). Finally, our findings have been validated in cohorts of individuals with both European and African ancestry. The vast majority of studies to date investigate solely individuals of European ancestry, while knowledge in non-European populations is very limited. Our study aimed to improve to knowledge on immune response regulation in both European and African populations. To achieve this aim, we have also built an online tool that can be used to explore efficiently our findings as an important research and development resource.

While our study provides extensive data on the cohorts and offers valuable insights, it still has some potential limitations. Firstly, confounding factors such as age, lifestyle, and genetic background, dietary habits (*52, 53*) can influence individual blood metabolite profiles. However, the consistent negative association between the sphingolipid metabolism pathway and cytokine responses across different cohorts and populations emphasizes its role in immune function and reassures on the validity of our conclusions. Secondly, while we employed the same mass spectrometry platform (flow-injection TOF-M) to measure metabolites across cohorts, this technique cannot precisely identify the isomers of metabolites. Since very long-chain SM and long-chain SM in plasma can play different roles in modulating inflammation (*54*), we also observed that the concentration of some SMs did not show a similar declining trend with the severity of COVID-19 as the majority of SMs (**fig. S9**). Thus, future research might delve deeper into the roles of different SM isomers in infectious diseases. In summary, this study provides valuable perspectives on the interplay between sphingolipid metabolism and immune responses in infectious diseases, further studies are needed to validate and expand upon these findings.

## Conclusion

Our study describes the interplay between metabolic signatures and immune functions across different cohorts and ethnic backgrounds, highlighting the potential of using metabolites as immunological modulators. The associations between metabolite-cytokine production revealed in this study are accessible for future research through an online tool known as IMetaboMap. Particularly, we identified that sphingolipid metabolism was associated with cytokine responses and disease risk in a severe infection (COVID-19). Sphingomyelins can mitigate inflammatory responses, providing tantalizing suggestions that diets rich in sphingomyelin may be useful for modulation of inflammation in immune-based diseases.

## Materials and Methods

### Cohorts’ descriptions

The first cohort, designated as Cohort_EU1, is part of the Human Functional Genomics Project as 500FG cohort and comprises 534 healthy Caucasian individuals of Western European ancestry, with ages spanning from 18 to 75 years. This cohort was carefully selected to exclude individuals with mixed genetic backgrounds or chronic diseases. Key measurements within Cohort_EU1 included cytokine production in response to various stimulations and comprehensive metabolomic profiling. More detailed information can be found in previous publications (*55*). The second cohort from Western Europe, Cohort_EU2, included 324 healthy volunteers of Western European descent, aged between 18 to 71 years. These participants were enrolled in the 300BCG cohort from April 2017 to June 2018, with further details documented in previous publication (*56*). The third cohort, Cohort_AF, encompasses 323 healthy Tanzanians between 18 to 65 years old from the Kilimanjaro region, who were recruited through the Kilimanjaro Christian Medical Center and Lucy Lameck Research Center from March to December 2017, as previously described (*20*).

### Plasma metabolome measurement and analysis

Untargeted metabolomics measurements from plasma were performed by high-throughput flow injection-time-of-flight mass spectrometry (*57*). The platform employed an Agilent 6520 Series Quadrupole Time-of-flight mass spectrometer and Agilent Series 1100 LC pump coupled to a Gerstel MPS2 autosampler. The metabolites were matched and annotated with HMDB (www.hmdb.ca), KEGG (www.genome.jp/kegg/) and ChEBI (www.ebi.ac.uk/chebi/) identifiers.

The NOREVA platform (http://idrblab.cn/noreva/) was employed to perform comprehensive data analysis using the peak intensity table of annotated metabolites (*58, 59*). Log transformation and Pareto scaling were applied before data analysis. Pathway enrichment analysis of identified metabolite lists was performed using the pathway analysis (Hypergeometric test) function of MetaboAnalyst V5.0 (https://www.metaboanalyst.ca/) (*60*). KEGG library was selected as the reference pathway library.

### Whole blood (WB) or peripheral blood mononuclear cells (PBMCs) stimulations

For WB stimulations, 100μl of heparin blood was diluted 1/5 with culture medium containing stimuli in 48-well plates and incubated for 48 hours with 100 ng/ml *E. coli*-derived *LPS*, 50 µg/mL Poly(I:C), 10^6^/mL *C. albicans*, 10^6^/mL *S. aureus*, 5 µg/mL *M. tuberculosis* (MTB), 10^6^/mL *E. coli*, 10^7^/ml *C. burnetii*, 10^7^/ml *S. pneumonia*, 10^6^/mL *S. typhimurium* or 10^6^/mL *S. enteritidis*.

5×105 PBMCs per well were stimulated in round-bottom 96-well plates with 5 µg/mL *M. tuberculosis* (MTB) or 106/mL *S. aureus* for 24 hours or 7 days. RPMI 1640 Medium (Dutch modification, Gibco) supplemented with 1 mM sodium pyruvate (Gibco), 2 mM GlutaMAX (Gibco), and 5 µg/ml gentamicin (Centraform) was used in all cell culture experiments. Supernatants were collected and stored at −20°C until cytokine quantification by ELISA. Cytokines were quantified using DuoSet ELISA Development Systems (R&D), except for the IFN-γ ELISA (Sanquin), according to the manufacturer’s instructions.

### Measurement of circulating cytokines concentrations

In the African cohort, cytokine concentrations in plasma were measured with the Ella platform (Protein Simple) using Simple Plex cartridges following the manufacturer’s protocols. In the European cohort, measurement of biomarker concentrations was performed using the Olink Inflammation Panel consisting of 92 markers (Olink Biosciences). This method employs proximity extension assay and provides relative protein quantification expressed as normalized protein expression (NPX) values (*61*).

### Statistical analyses

To elucidate the metabolic networks underlying immune phenotypes, we assessed the metabolite co-expression networks of immune responses (IL-1β, IL-6, TNF, IFN-γ) following *S. aureus* stimulation. Utilizing Weighted Correlation Network Analysis (WGCNA) (*62*), we identified modules of metabolites exhibiting highly correlations (fig. S1). We proceeded to examine the relationship between the summary profile (eigengene) of each module and various different immune responses (IL-1β, IL-6, TNF, and IFN-γ), as depicted in Fig. 2. Upon discovering any modules with significant correlations to immune responses, we isolated the metabolites within these modules for further exploration. Pathway analyses were conducted using MetaboAnalyst 5.0. We reported metabolic pathways achieving a threshold of p < 0.05, which are presented.

We computed the correlation between individual metabolite features and the immune responses to *S. aureus* across multiple cohorts. Among 3 cohorts, a high proportion of metabolites was significantly correlated with sex (Cohort_AF: 38.4%, Cohort_EU1 (PBMCs and WB): 50.5%, Cohort_EU2: 38.9%), age (Cohort_AF: 45.2%, Cohort_EU1 (PBMCs and WB): 36.8%, Cohort_EU2: 15.4%), and BMI (Cohort_AF: 16.9%, Cohort_EU1 (PBMCs and WB): 15.7%, Cohort_EU2: 0.51%). After accounting for age and sex, a smaller proportion of metabolites maintained their significant association with BMI (Cohort_AF: 6.02%, Cohort_EU1 (PBMCs and WB): 9.73%, Cohort_EU2: 0.07%, FDR <0.05,). Therefore, in the correlation analysis described below, we adjusted for the confounding effects of age and sex. To calculate the relationship between metabolites and immune cytokines, we first filtered features based on their Spearman correlation p-values, retaining only those features that passed specific thresholds (0.05 for metabolites) for further analysis. Details of the method can be found in a previous paper (*16*).

Apart from the MetaboAnalyst v.5.0 platform, statistical analyses were performed using R 4.1.0 (www.R-project.org). Correlation analysis was computed by the R packages ‘corrplot’ or ‘gplots’ upon Spearman’s rank correlation and Benjamini-Hochberg correction using the ‘corr.test’ function. We adjusted the effect of age, sex, and BMI on metabolite levels using a linear regression model, and calculated false discovery rate (FDR < 0.05).

### Meta-analysis

To prepare for the meta-analysis, we first converted Spearman’s correlation coefficients into Fisher’s Z values. This transformation is a crucial step that stabilizes the variances and normalizes the distribution of the coefficients, which is essential for the subsequent pooling of data across different studies. For each Fisher’s Z value, we calculated its standard error (SE) using the formula

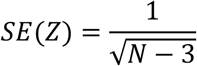

where N represents the sample size from the respective study.

Our meta-analytical approach consisted of both fixed-effect and random-effect models, allowing us to explore the consistency of the effect sizes across studies and account for any between-study heterogeneity. The fixed-effects model was implemented under the assumption that the effect sizes are homogenous, while the random-effects model, incorporating the REML method, was employed when heterogeneity was present. This approach acknowledges that individual studies might estimate different, yet related, effects and thus incorporates both within-study and between-study variation into the analysis. The ‘metafor’ package (*63*) facilitated the computation of pooled effect sizes and their corresponding 95% confidence intervals. We further generated forest plots to visually represent the individual study effects, their confidence intervals, and the pooled estimate, providing a clear and concise graphical summary of the meta-analysis results.

### Mendelian randomization

MR analysis was done using the TwoSampleMR (version 0.5.6) R package. The main two-sample MR methods used in this study include IVW (*37*) and weighted median (*64*). Therefore, MR estimates were calculated using Wald ratios and these Wald ratios were meta-analyzed using the IVW method (*37*). To ensure the validity of the results, several sensitivity analyses were performed. We excluded MR estimates potentially driven by horizontal pleiotropy (removing results with MR-Egger (*38*) intercept P < 0.05) and heterogeneity (removing results with Cohran’s Q test P < 0.05). In addition, we carried out leave-one-out analysis (*65*) to check whether the MR estimates were possibly driven by a single SNP. Multiple testing correction was performed using the Benjamini–Hochberg approach based on IVW P values. To avoid complex causality relationships, we excluded the results that showed a nominally significant MR estimate in the other direction (P < 0.05). For this analysis, metabolite-associated SNPs at a P value cut-off of 5 × 10^−7^ used as genetic instruments in IVW-based MR. Due to the limited number of participants in the study and our aim to identify a broader range of potentially significant SNPs for subsequent MR analysis, we have chosen a more relaxed yet still conservative significance threshold of 5 × 10^−7^. This approach is informed by similar strategies in genetic research, where such a threshold has been employed to balance the trade-off between sensitivity and specificity, particularly in contexts where maximizing the discovery of candidate SNPs is crucial(*66–68*).

### In vitro validation experiments

Venous blood was sampled in sterile 10mL ethylenediaminetetraacetic acid (EDTA) tubes (Vacutainer system, Becton Dickinson) from six healthy volunteers (ethical approval NL84281.091.23 of the Arnhem-Nijmegen Ethical Committee). Peripheral blood was processed within 1-4 hours after collection. Human PBMCs were isolated by differential centrifugation by Ficol-Paque, as previously described (*69*). PBMCs were stimulated with either lipopolysaccharide (LPS from E. coli 055:B5, Sigma) or heat-killed Staphylococcus aureus for 24 hours, in the absence or presence of different concentrations of chicken yolk or porcine brain sphingomyelin (10 ng/ml to 10 ug/ml). Culture supernatants were collected at the end of incubation period and stored at −20°C until cytokines were measured using enzyme-linked immunosorbent assay. The production of proinflammatory cytokines was measured using commercial ELISA kits (R&D Duoset ELISA Systems) according to the manufacturer’s instructions.

### Visualization

R package ggplot2 was used to perform most visualizations, including correlation plots, bar charts, and boxplots.

### Online tool implementation details

IMetaboMap was developed by R v 4.3.2 and Shiny v 0.13.1 running on Shiny-server v1.7.5. Various R packages were utilized in the background processes. IMetaboMap can be readily accessed by all users with no login requirement, and by diverse and popular web browsers including Google Chrome, Mozilla Firefox, Safari and Internet Explorer 10 (or later).

## Data Availability

All data produced in the present work are contained in the manuscript

## Acknowledgments

The authors thank all volunteers from the Radboud University Medical Center and Tanzania for their participation in the study. This work was supported with funds provided by ERC starting grant 948207, I ASPASIA, and Radboud University Medical Center Hypatia grant to Y.L., ERC advanced grant 833247 and Spinoza grant of the Netherlands Organization for Scientific research to M.G.N., and Helmholtz Initiative and Networking Fund to C.J.X. (1800167). This study was also supported by the COFONI (COVID19 Research Network of the State of Lower Saxony) with funding from the Ministry of Science and Culture of Lower Saxony, Germany (14-76403-184-3) and the Deutsche Forschungsgemeinschaft (DFG; German Research Foundation) under Germany’s Excellence Strategy—EXC 2155 project number 390874280 to Y.L. The study received financial support from the Joint Programming Initiative, A Healthy Diet for a Healthy Life (JPI-HDHL), and ZonMw (the Netherlands Organization for Health Research and Development), within the framework of the ‘TransMic’ and ‘TransInf’ projects.

## Author Contributions

Conceptualization, Y.L., J.F.; formal analysis, J.F.; investigation, N.N., N.V.U.; resources, M.J., S.M., V.K., C.B., V.M., G.T., V.K., Q.M., and L.A.B.J.; validation, A.S. and M.G.N.; supervision, Y.L. and M.G.N.; writing original draft, J.F., N.N., Y.L., C. X. and M.G.N.; writing– review & editing, all authors.

