## Supplemental Figures for "Deciphering Cross-Cohort Metabolic Signatures of Immune Responses and Their Implications for Disease Pathogenesis"

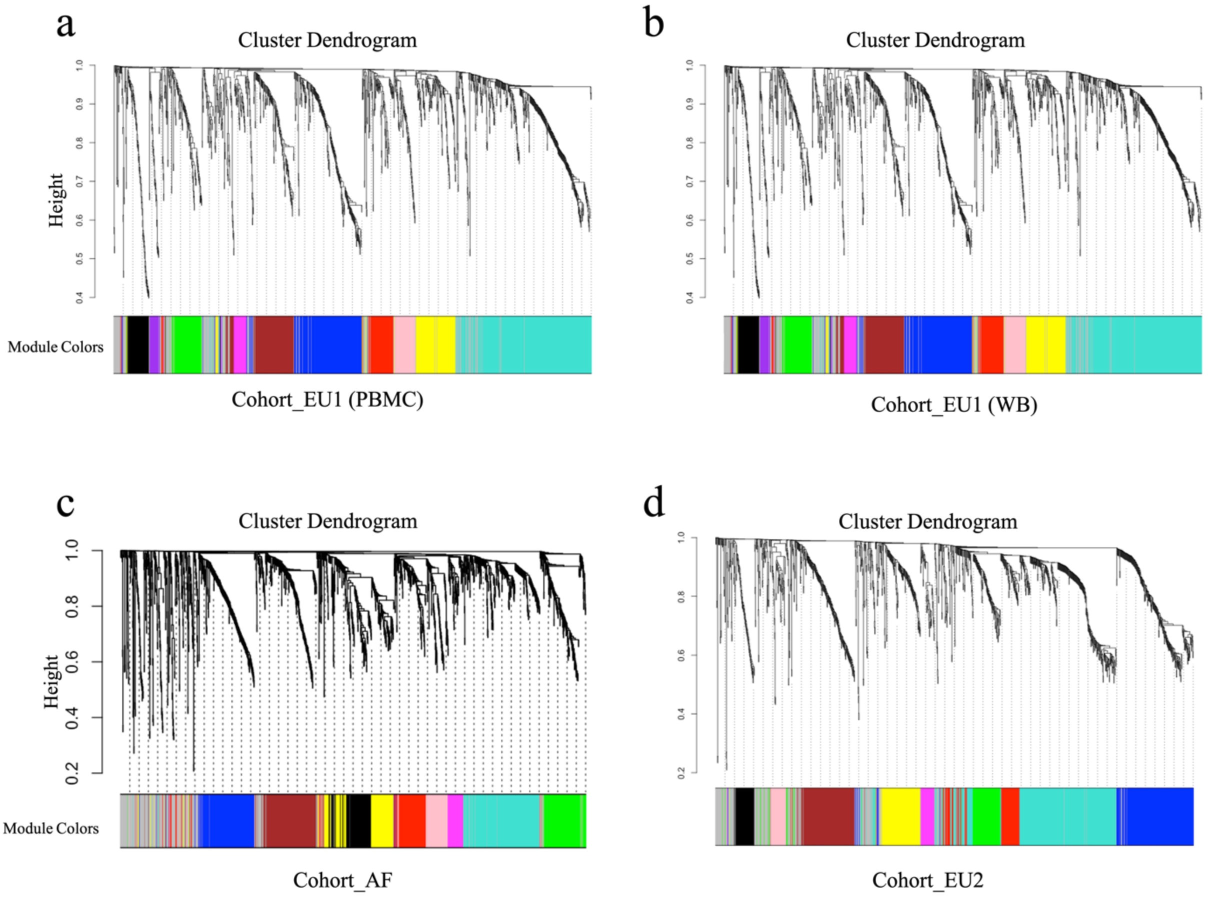


**Fig. S1. Hierarchical clustering dendrogram of metabolites data (a, b and c).** The WGCNA analysis reveals the association of metabolite groups with cytokine responses (IL-1β, IL-6, TNF, and IFN-γ) triggered by S. aureus. The specific metabolites within each module delineated by WGCNA are detailed in the Table S1.

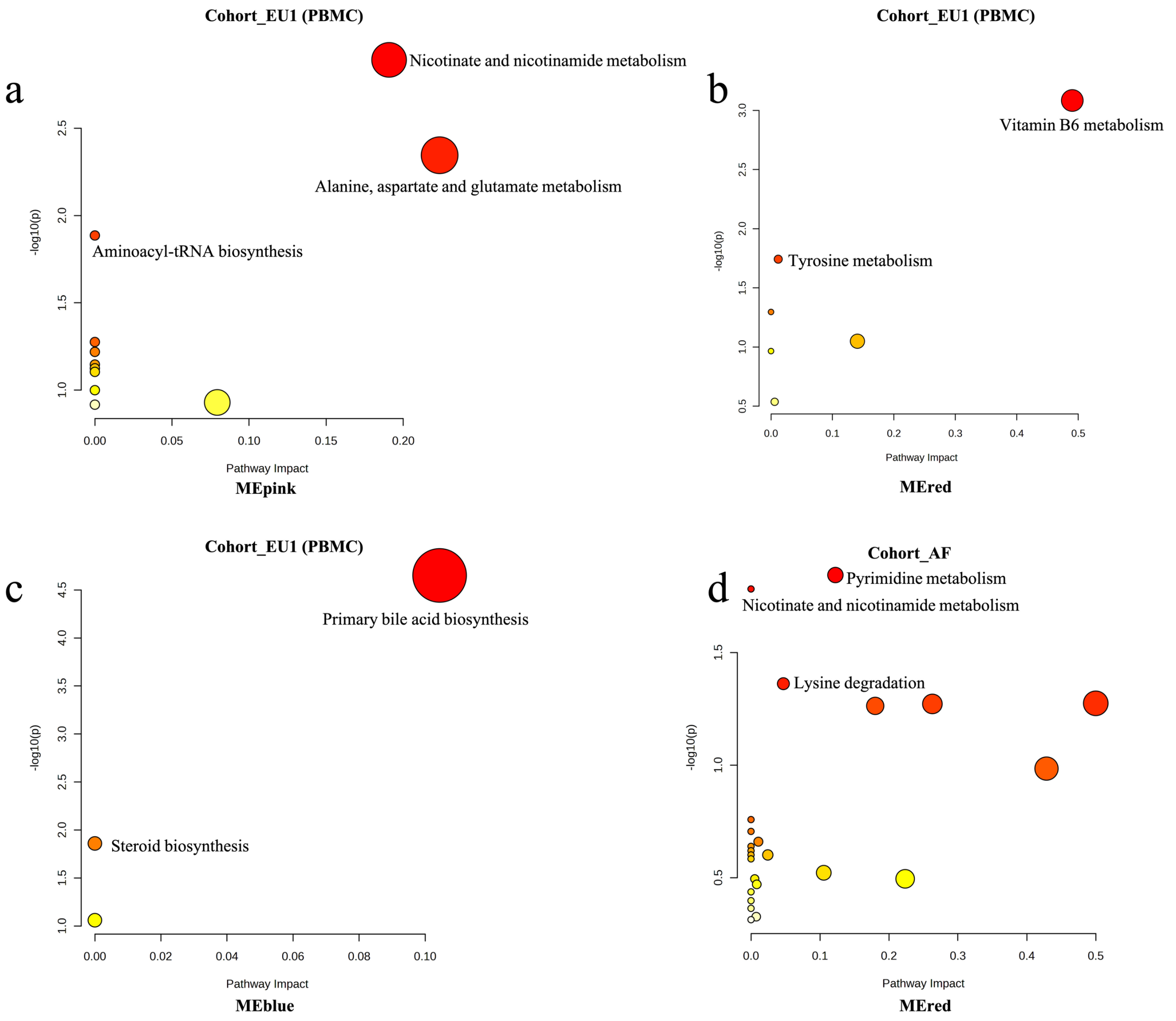


**Fig. S2. Pathway analysis results for metabolites modules within the Cohort_EU1 and Cohort_AF.** Scatter plots illustrate the pathway analysis results for metabolites within the MEpink (Fig. 2a), MEred (Fig. 2a), MEblue (Fig. 2a) modules of Cohort_EU1 and MEred (Fig. 2b) module of Cohort_AF. The darker the bubble color, the larger the -log(Pvalue), indicating higher significance.

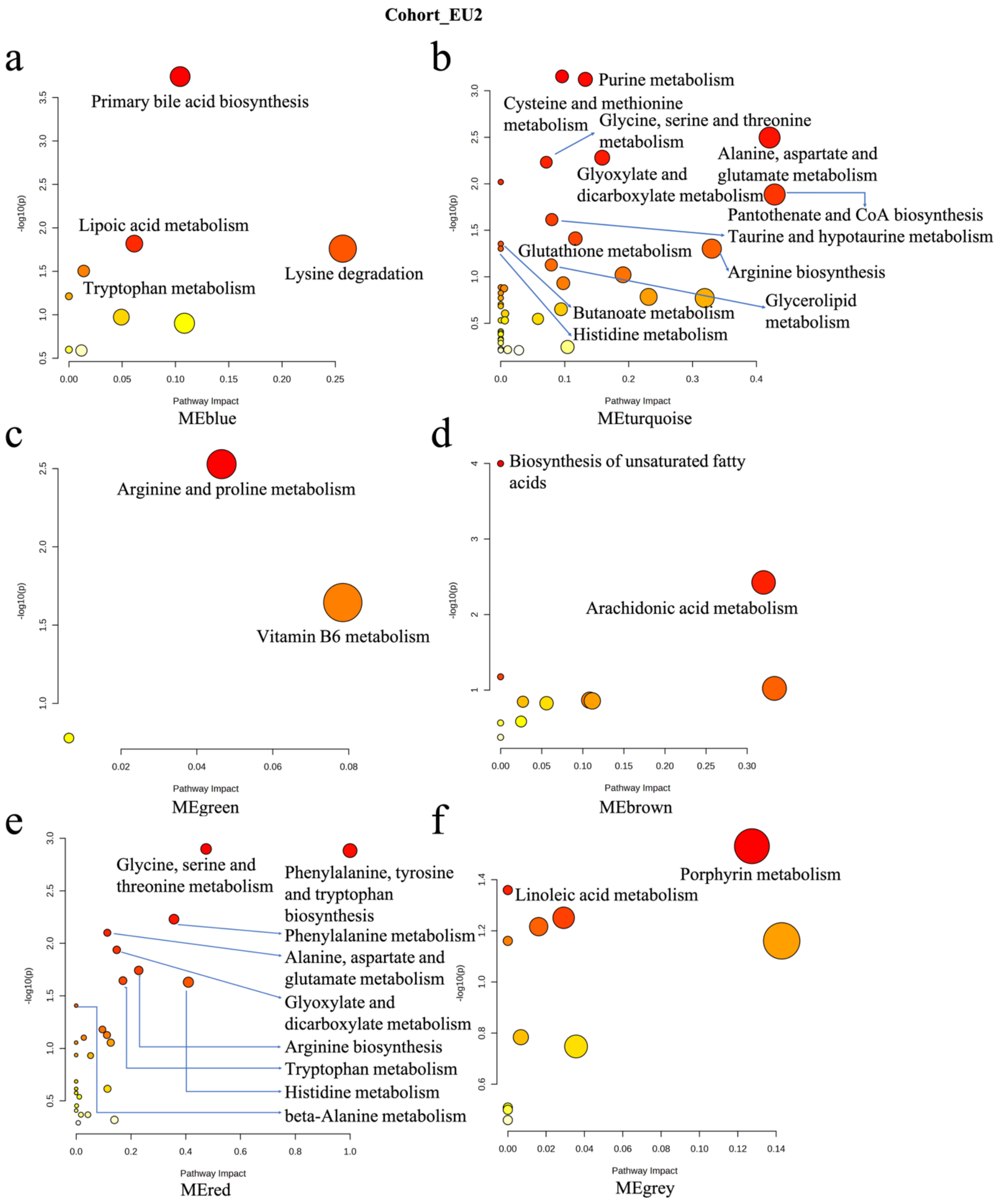


**Fig. S3**. **Pathway analysis results for metabolites modules within the Cohort_EU2.** (a-f) Pathway analysis results for metabolites within the MEblue, MEgreen, MEred, MEturquoise, MEbrown, and MEgrey modules of Fig. 2d. The darker color and larger size of the bubble dots indicate a larger -log(Pvalue), suggesting higher significance.

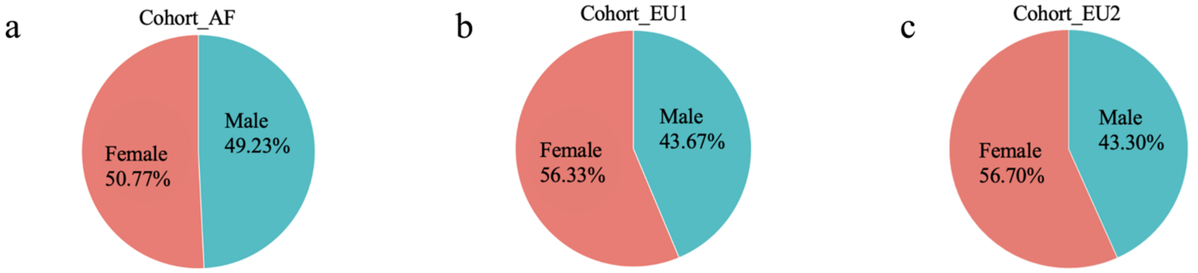


d

| Cohort | Cohort_AF | | Cohort_EU1 | | | | Cohort_EU2 | |
|---|---|---|---|---|---|---|---|---|
| Tissue | WB | | PBMCs | | WB | | PBMCs | |
| Gender | Male | Female | Male | Female | Male | Female | Male | Female |
| Total metabolites (fdr ≤ 0.05) | 59 | 10 | 95 | 302 | 7 | 52 | 0 | 0 |
| Total metabolites (0.05< fdr < 0.1) | 121 | 20 | 97 | 139 | 16 | 87 | 0 | 0 |

**Fig. S4**. **Sex differences impact the correlation between metabolites and cytokine responses.** (a) Cohort_AF, Cohort_EU1 and Cohort_EU2 populations with the distribution of sex. (b) The number of metabolites associated with the correlation between S. aureus stimulus-induced cytokine responses (IL-1β, IL-6, TNF, and IFN-γ) in males and females from the different cohorts was illustrated.

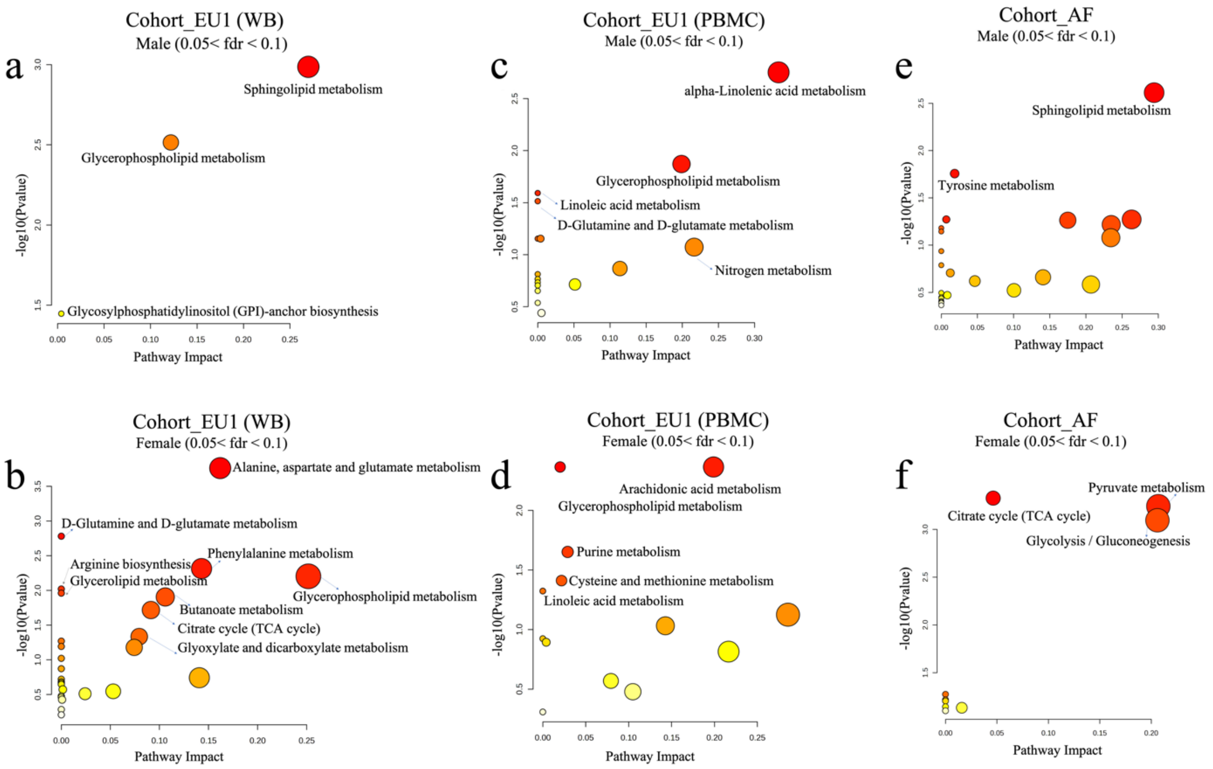


**Fig. S5**. **Scatter plots illustrate the results of the pathway analysis** obtained by correlating males and females separately in different cohorts with correlated metabolites (0.05 < FDR < 0.1) with either cytokine responses (IL-1β, IL-6, TNF, and IFN-γ) induced by S. aureus stimulation. The darker the bubble color, the larger the -log(Pvalue), indicating higher significance.

a

| Cohort | Cohort_AF | Cohort_EU1 | | Cohort_EU2 |
|---|---|---|---|---|
| Tissue | WB | PBMCs | WB | PBMCs |
| Total metabolites (fdr ≤ 0.05) | 255 | 423 | 167 | 0 |
| Total metabolites (0.05< fdr < 0.1) | 210 | 223 | 138 | 0 |

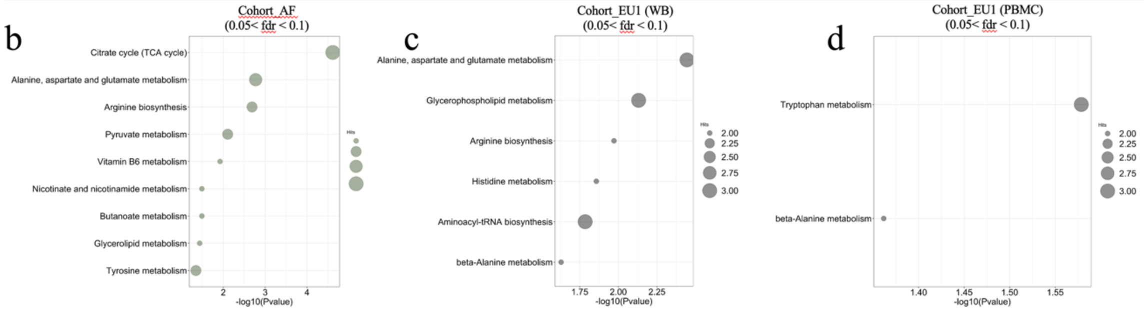


**Fig. S6**. **Immune response-related metabolites across multiple cohorts.** (a) The number of metabolites associated with the correlation between S. aureus stimulus-induced cytokine responses (IL-1β, IL-6, TNF, and IFN-γ) in the different cohorts is illustrated. (b, c and d) Bubble plots showing the results of pathway analysis of metabolites significantly correlated (0.05 < FDR < 0.1) with S. aureus induced cytokine responses (IL-1β, IL-6, TNF, and IFN-γ) in different cohorts.

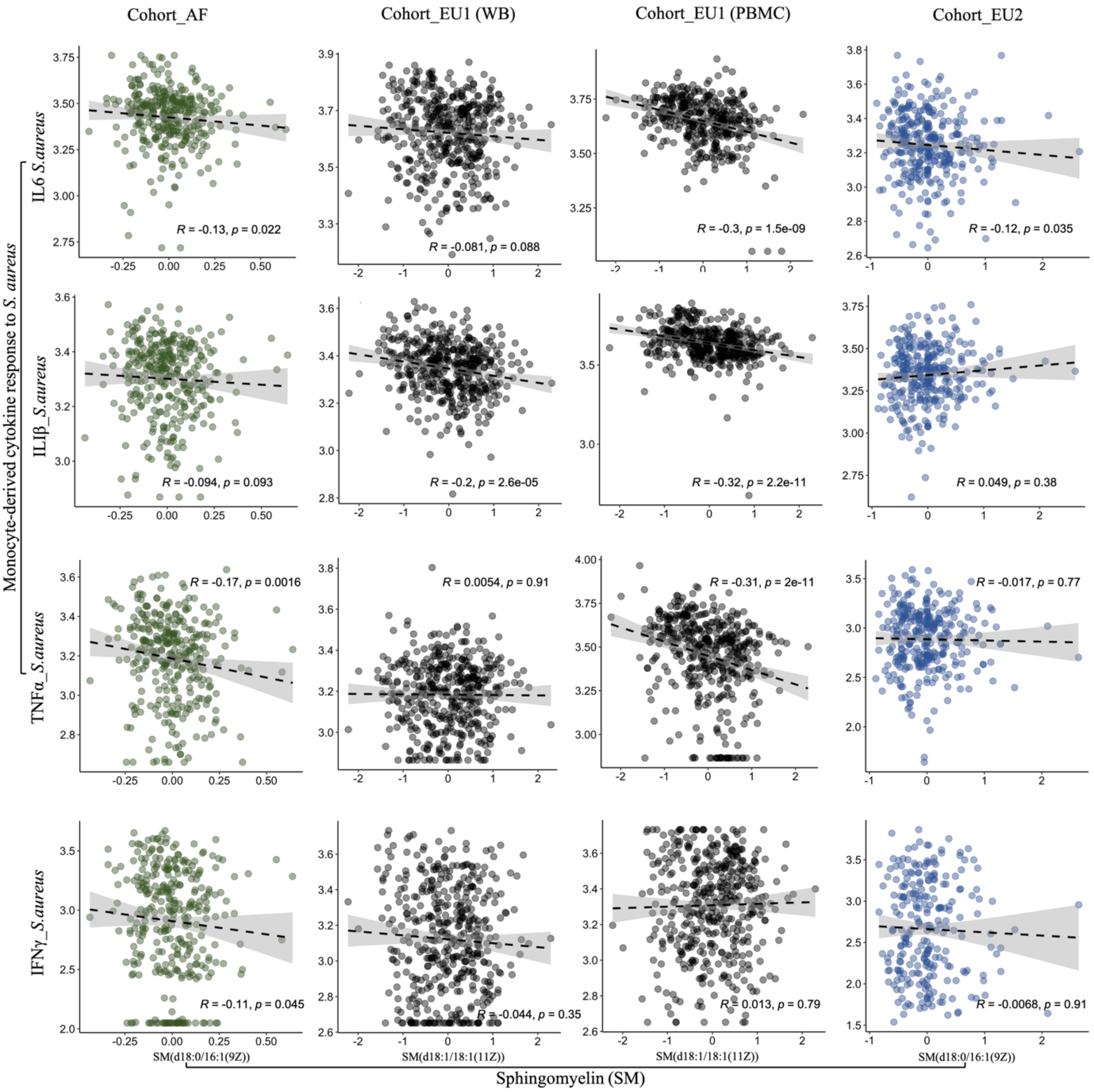


**Fig. S7**. **Linear regression** between sphingomyelin (SM) at baseline and S. aureus induced cytokine responses (IL-1β, IL-6, TNF, and IFN-γ) in the Cohort_AF, Cohort_EU1 and Cohort_EU2. r: Spearman's correlation coefficient.

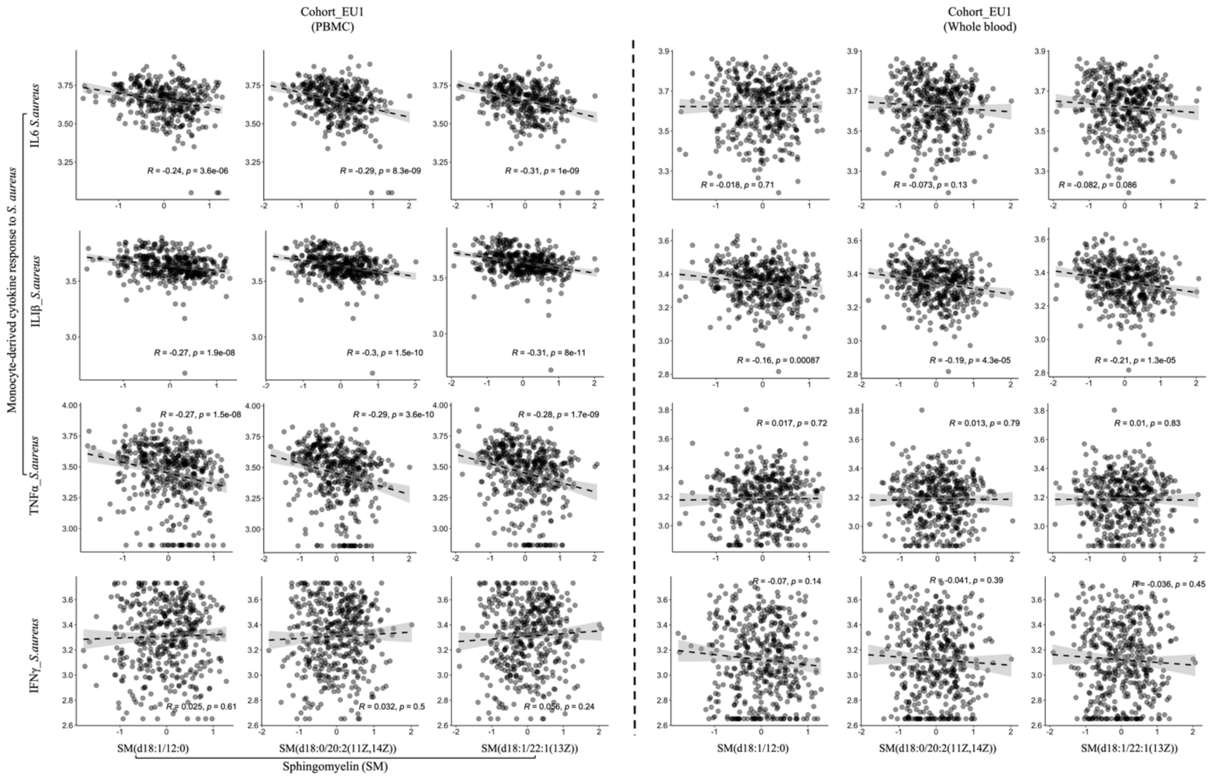


**Fig. S8**. **Linear regression** between sphingomyelin (SM) at baseline and S. aureus induced cytokine responses (IL-1β, IL-6, TNF, and IFN-γ) in the Cohort_EU1. r: Spearman's correlation coefficient.

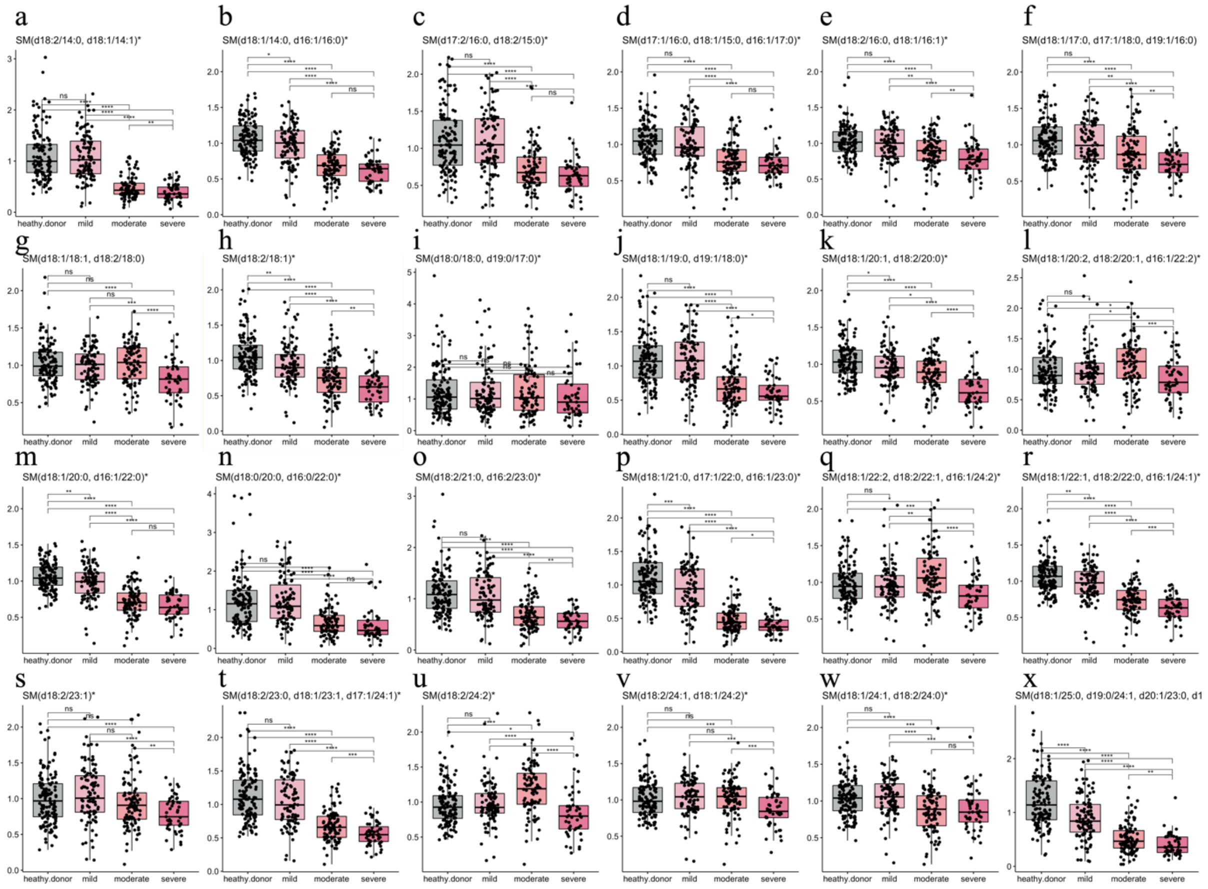


**Fig. S9**. **Changes in sphingomyelins of sphingolipid metabolism in COVID-19 patients.** (A-X) Gray denotes a healthy individual while increasing shades of red indicate escalating severity of COVID-19 (*p<0.05, **p<0.01, ***p<0.001).
